## Supplementary Information for "Fine scale human mobility changes in 26 US cities in 2020 in response to the COVID-19 pandemic were associated with distance and income"

### Supplementary material

1. Modification to Poisson likelihood for censored observations

First, consider the model of decreasing mobility between February 2 – April 4. The number of trips between zip codes $i$ and $j$ (in either direction) in age group $a$ at time $t$ is denoted $Y_{ijat}$. For simplicity we can considered a fixed time and age group and just write $Y_{ij}$ for this variable. This is the sum of trips from $i$ to $j$ and $j$ to $i$, which we denote $Y_{i-j}$ and $Y_{j-i}$ respectively.

Let $\lambda$ be the mean of the Poisson process implied by some parameter values (again just for notational convenience), so the likelihood when $Y_{ij}$ is known exactly is

$$Y_{ij} \sim Poisson(\lambda)$$

or in other words

$$p(Y_{ij}|\lambda)$$

where $p$ is the probability mass function of a Poisson distribution with mean $\lambda$.

There are two possibilities for when the total $Y_{ij}$ isn't known exactly, either one of $Y_{i-j}$ and $Y_{j-i}$ are below 50 (but not both) or both are below 50. Suppose just one is below 50 and let it be $Y_{i-j}$ be known (otherwise switch the indices). The total $Y_{ij}$ could therefore be any value between $Y_{i-j}$ and $Y_{i-j} + 49$ and so the likelihood is

$$p\left(Y_{i-j} \leq Y_{ij} \leq Y_{i-j} + 49 \middle| \lambda\right) = \sum_{k=0}^{49} p(Y_{i-j} + k|\lambda)$$

Similarly, if both are below 50 then $Y_{ij}$ could take any value between 0 and 99, yielding a likelihood

$$p\left(0 \leq Y_{ij} \leq 99 \middle| \lambda\right) = \sum_{k=0}^{99} p(k|\lambda)$$

Likelihoods were modified in a similar way for the model of mobility between June 1 – August 31, where the uncertainty (and therefore the number of values summed over) accumulates for each week that is censored.

### 2. Prior distributions

#### *Modelling initial decrease in mobility*

All effect sizes were given Normal priors centred at zero with standard deviations of 1. Baseline trip rates were parameterised on the log scale with Normal priors centred at -3 with standard deviations of 0.5. Note that baseline trip rates were treated as random parameters and integrated out (via a Laplace approximation) at each step of the optimisation. The weekly city-wide rates of decrease in mobility, $c_k$, were also parameterised on the log scale with an AR(1) prior, with $\phi = 1,\ c = 0, \epsilon = 0.1$.

#### *Modelling mobility over summer*

All effect sizes were given Normal priors centred at zero with standard deviations of 1.

Figure S1: Daily trips relative to baseline in cities in the South (excluding San Antonio) between June 1 – August 31, with days with data loss removed. Using daily trips reduces the amount of missingness as in the weeks with data loss there was only data loss in some (not all) days. With the exception of Phoenix, where travel decreased during June from above baseline levels to around 90% of baseline, there is little evidence of a decreasing mobility during this time frame. San Antonio is not included as there was a similar amount of data loss in weekly and daily data.

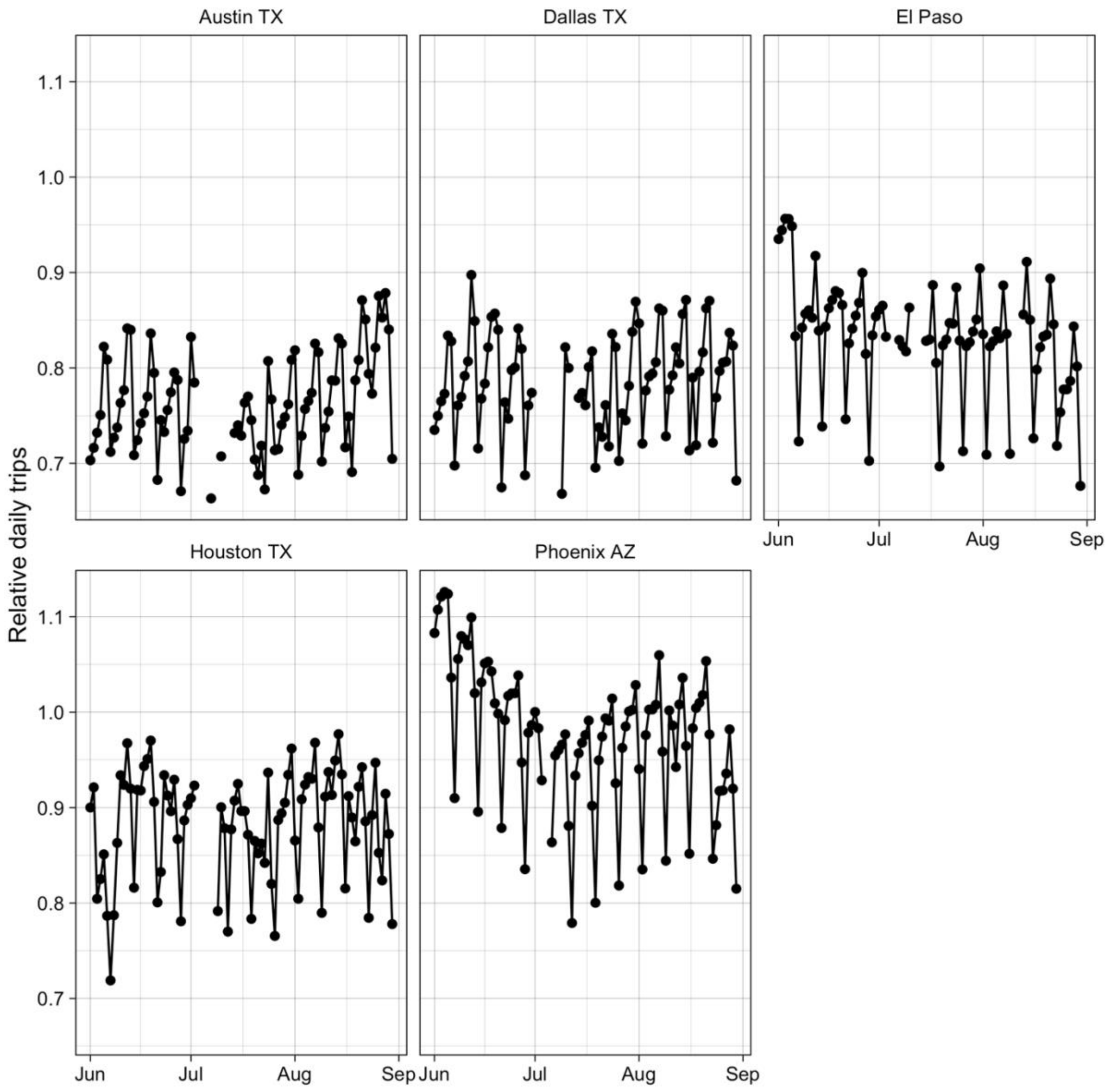

Table S1: Summary of all models used

| | Model 1: Rate of decrease in mobility | Model 2: Summer mobility compared to baseline | Model 3: Comparison to model 1 – rate of decrease in mobility |
|---|---|---|---|
| Time frame | February 1 – April 4 | June 1 – August 31 | February 1 – April 4 |
| Baseline component | Baseline rate of travel between each pair of zips is a parameter to be learned. | Baseline rate of travel between each pair of zips is fixed at the rate learned in model 1. | Gravity model where propensity to travel and effect of distance, origin population and destination population on baseline travel is learned. |
| Changing mobility component | For each week there is a city-wide parameter for average rate of decrease of mobility. The effect of each explanatory variable on this city-wide average is learned. | Parameter $\beta_0$ describes average change in mobility rates between baseline and summer. The effect of each explanatory variable on this city-wide average is learned. | Same as model 1 |
| Explanatory variables | Distance, proportion of high-income subscribers, case rate relative to city, age, median household income | Distance, proportion of high-income subscribers, age, median household income | Same as model 1 |

<u>3. List of zip codes used</u>

See file “list_of_zips.csv” (dataset 1)

<u>4. Weeks removed due to data loss</u>

See file “weeks_removed.csv” (dataset 2)

### 5. Trips by age

Figure S2: Trips relative to baseline by age group in each city (a) – (e)

Figure S2(a)

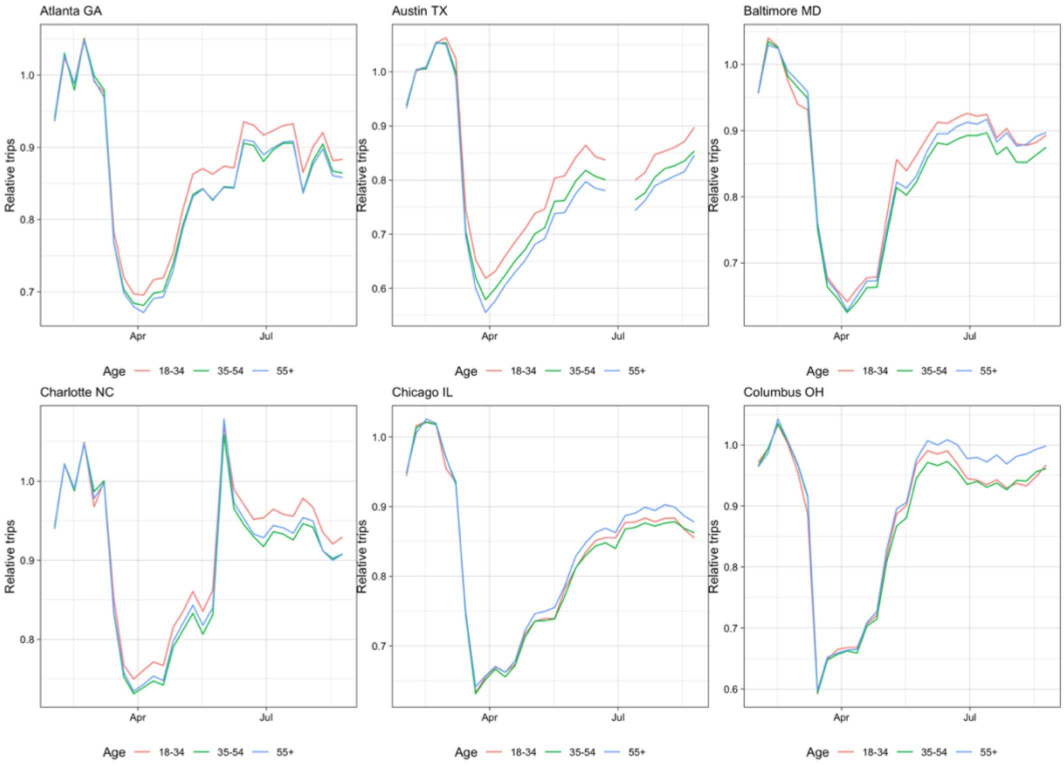


Figure S2(b)

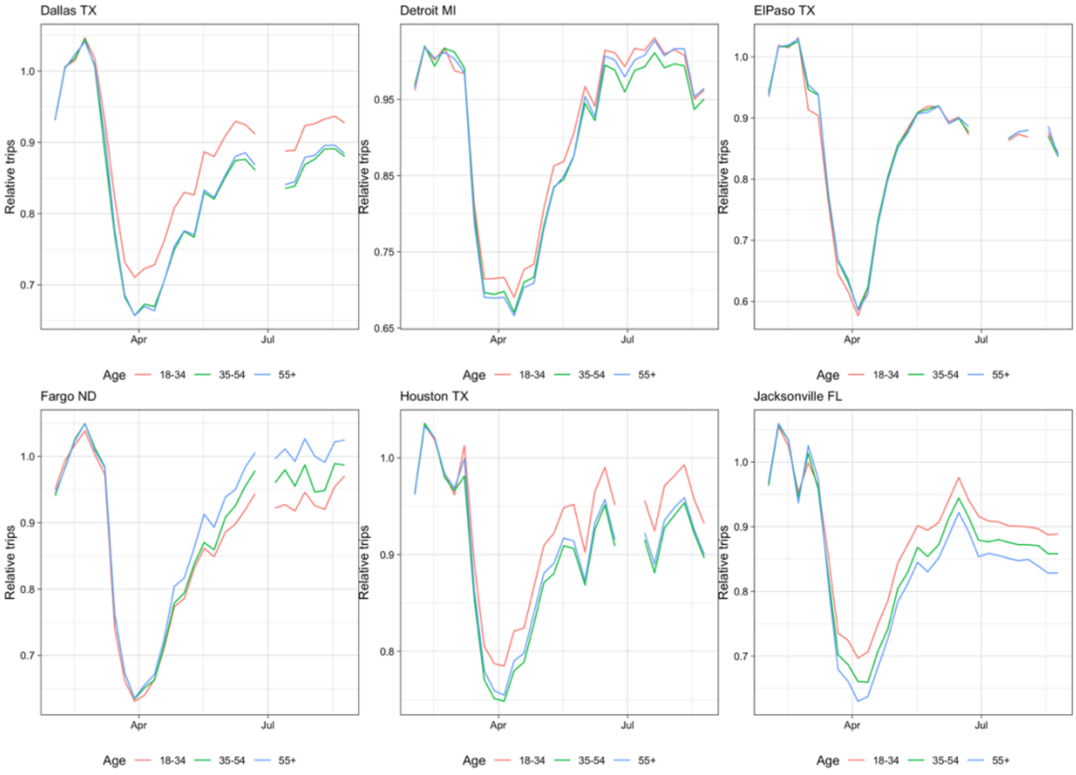

Figure S2(c)

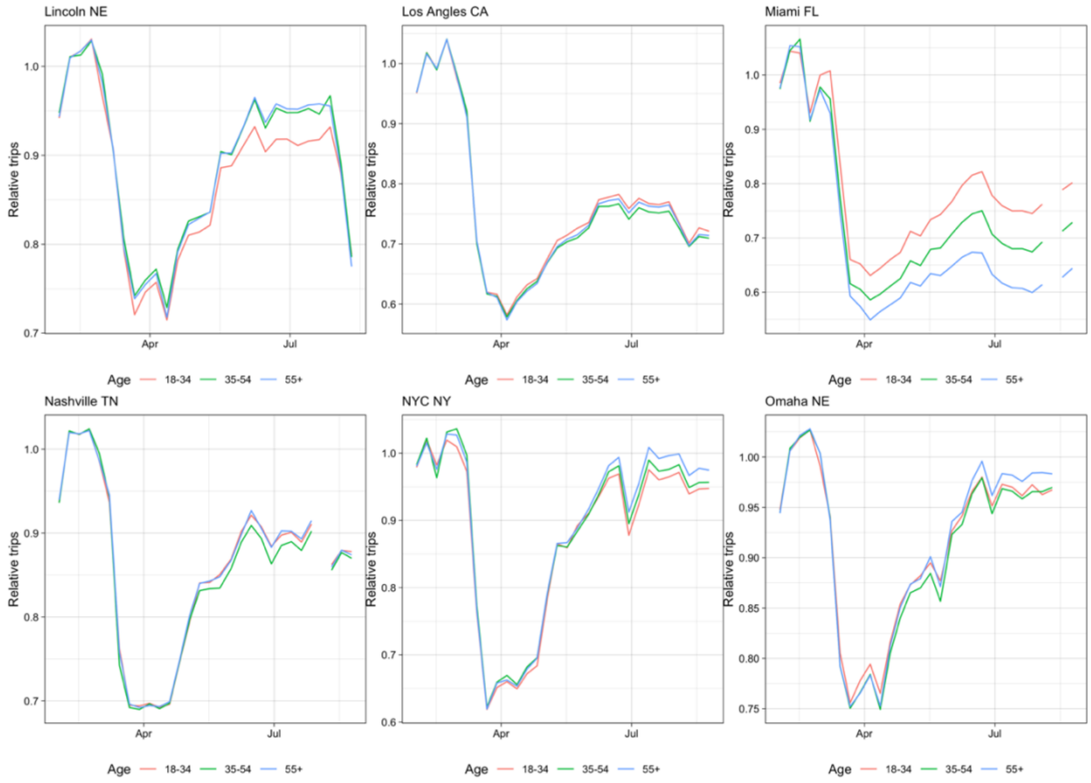


Figure S2(d)

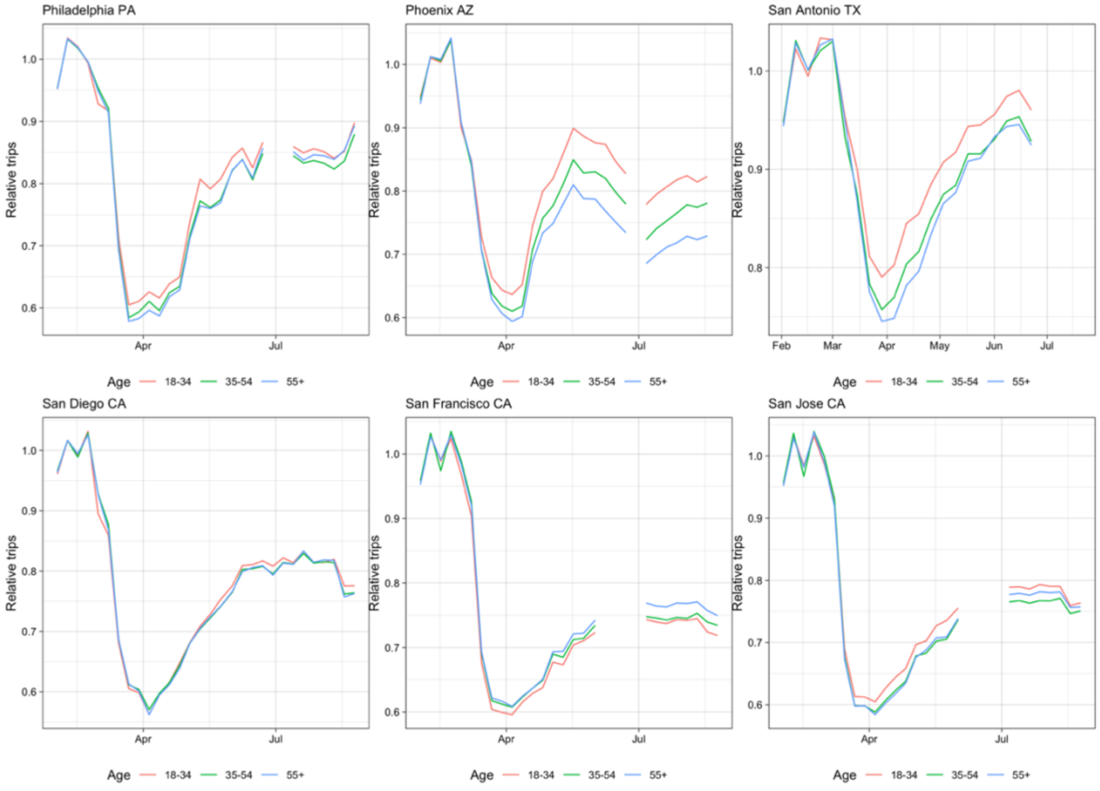

Figure S2(e)

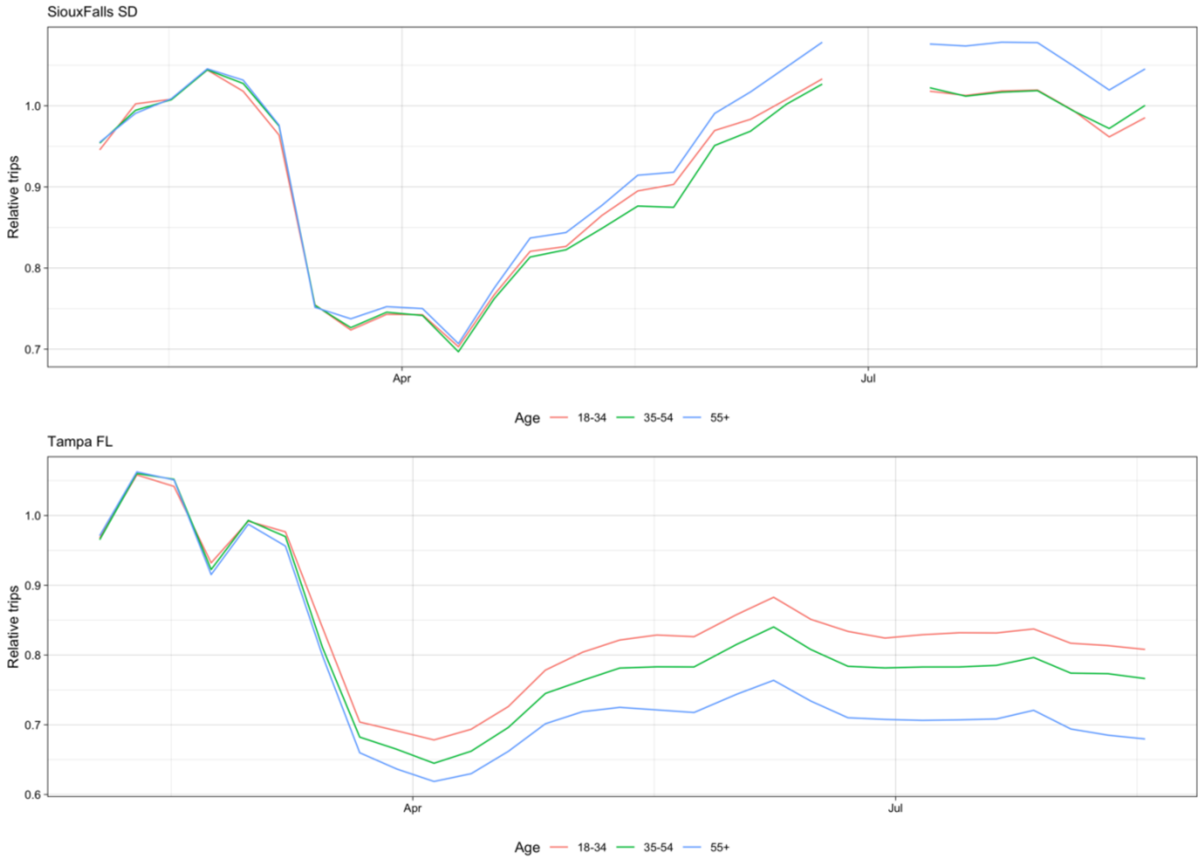
SiouxFalls SD
Relative trips
1.0
0.9
0.8
0.7
Apr
Jul
Age
18-34
35-54
55+
Tampa FL
Relative trips
1.0
0.9
0.8
0.7
0.6
Apr
Jul
Age
18-34
35-54
55+

<u>6. Trips by distance</u>

Figure S3: Relative trips by distance quartile for each city (not including those presenting in the main text) (a) – (d)

Figure S3(a)

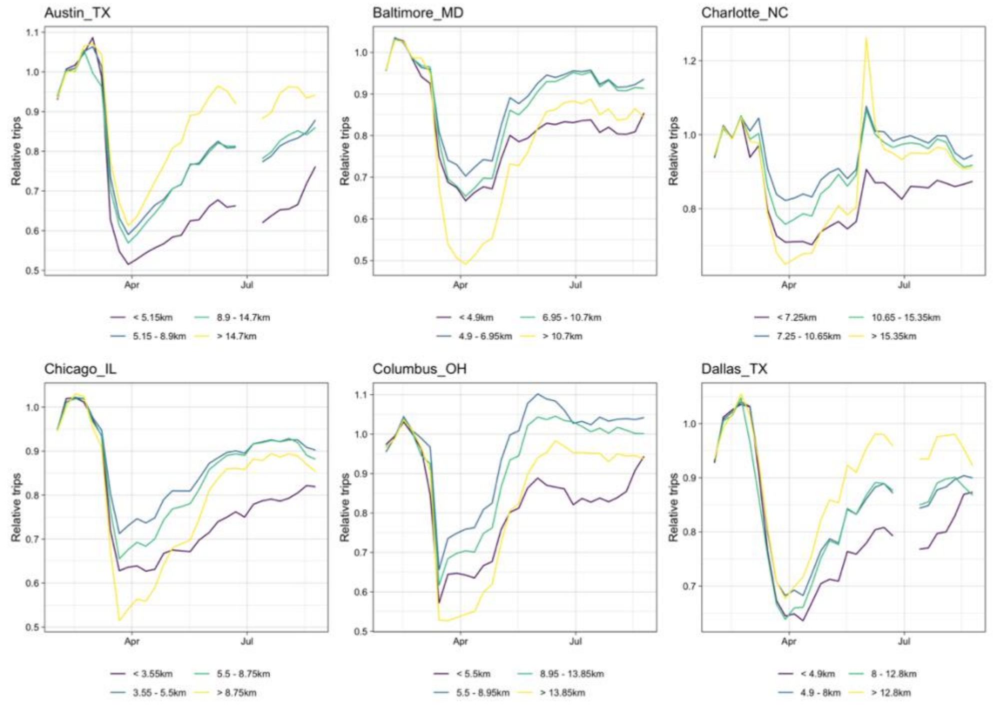


Figure S3(b)

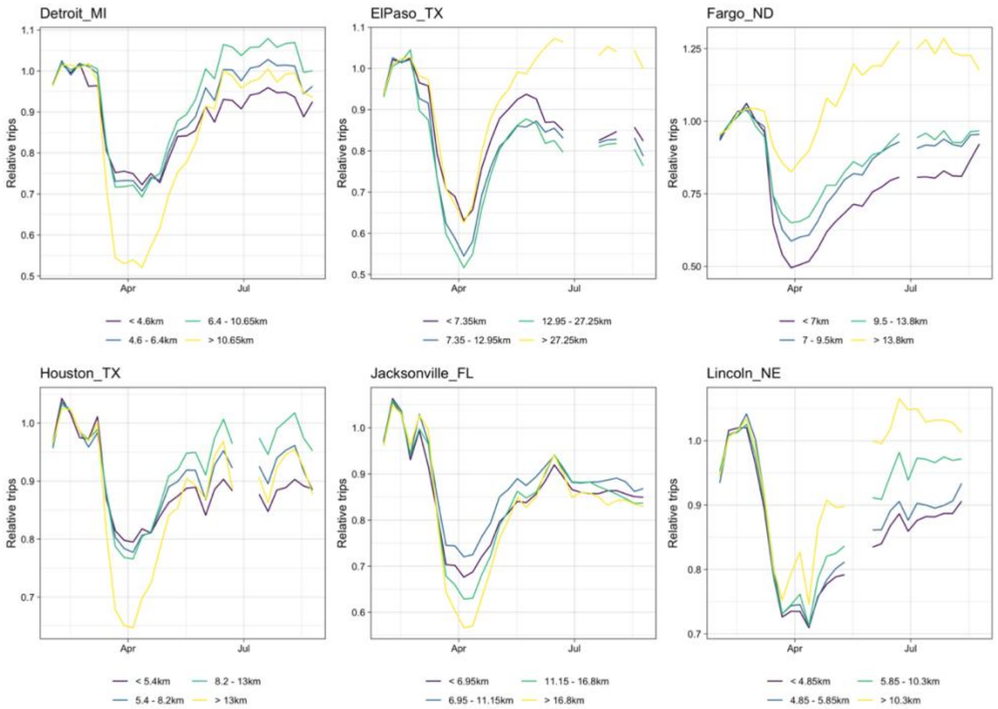

Figure S3(c)

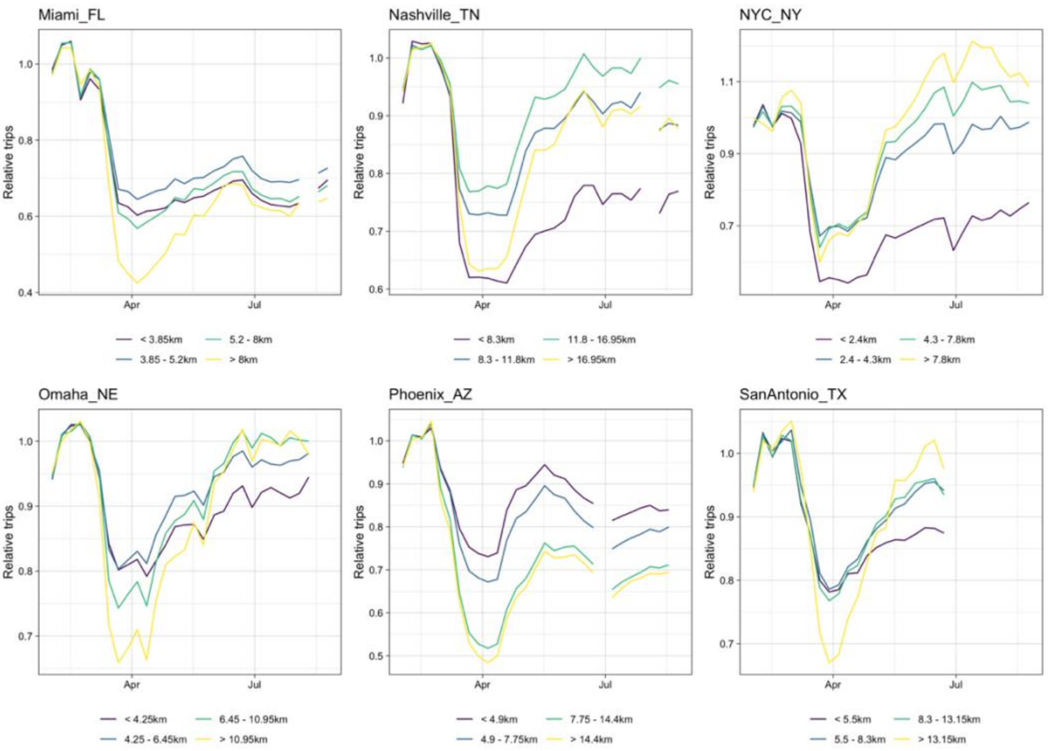


Figure S3(d)

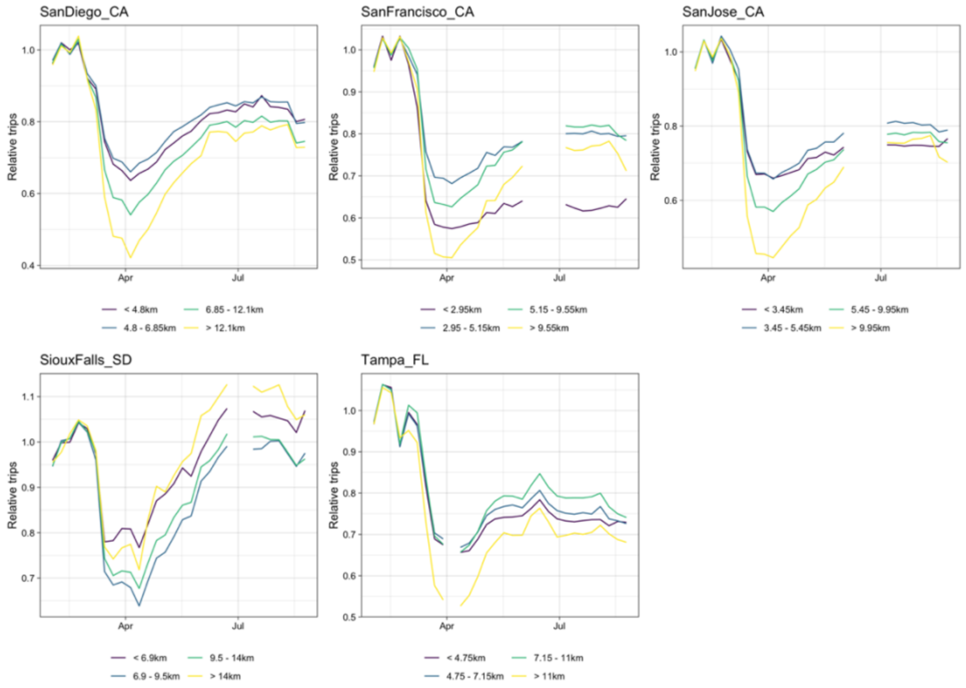

### 7. Demographic distributions

The distributions of individual income for the subscribers in the dataset in each city is shown in Figure S4 and for the general population of the US by region from Census data in Figure S5. A clear overrepresentation of higher income groups can be seen in all cities compared to the general population in any region.

Figure S4: Proportions of subscribers in each city in each income bracket

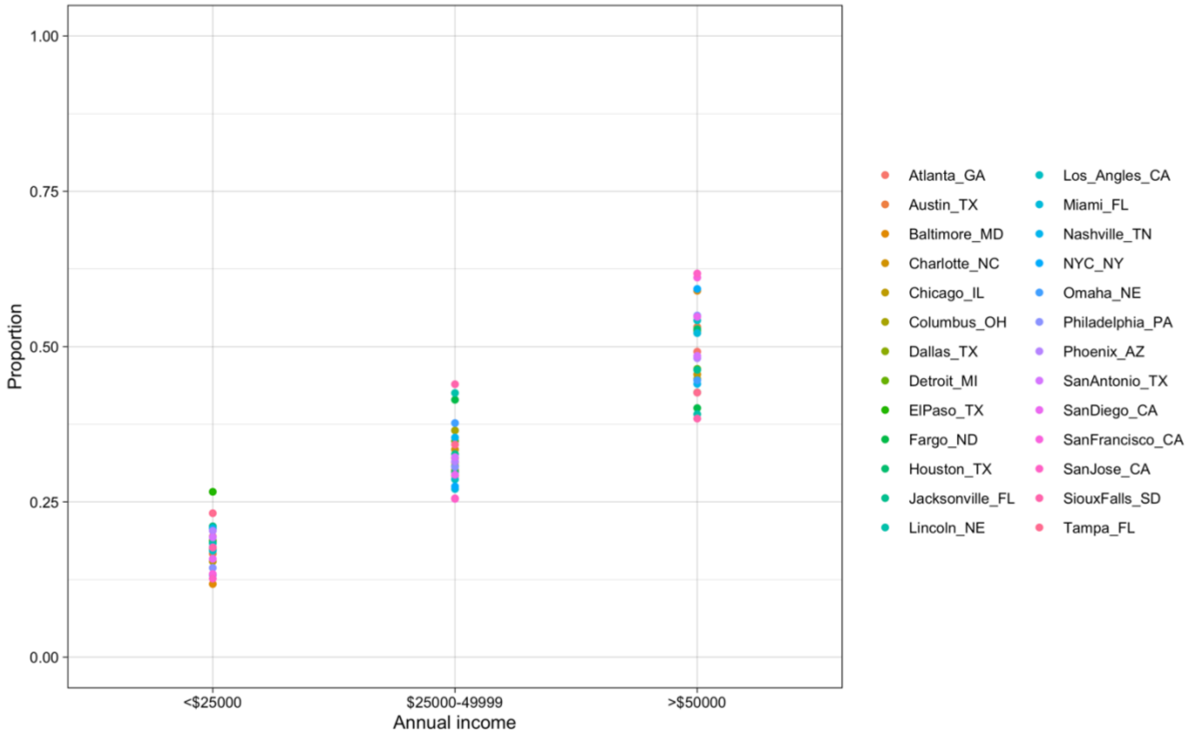

Figure S5: Proportion of general population in regions of the US in each income bracket [60]

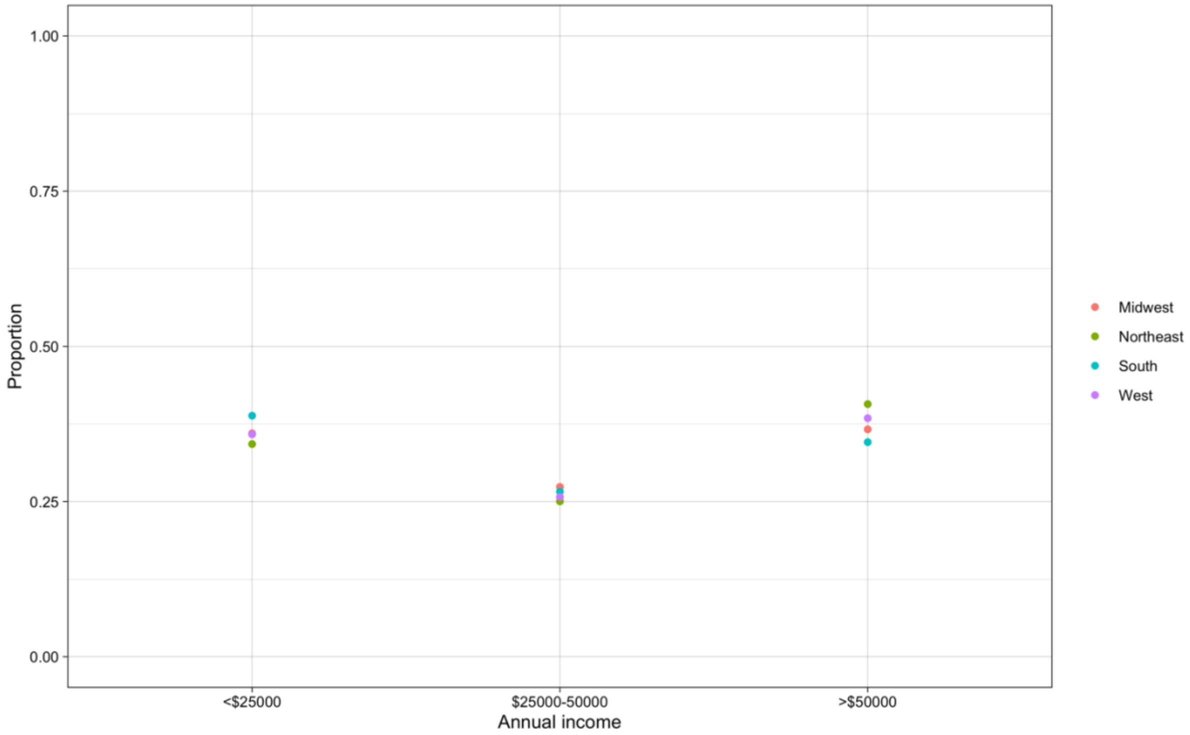


The distributions of age for the subscribers in the dataset in each city is shown in Figure S6 and for the general population of the US from Census data in Figure S7. Here the older age groups are overrepresented in the subscriber data compared to the general population, which is clear despite the slight mismatch in the age ranges of the youngest group.

Figure S6: Proportions of subscribers in each city in each age bracket

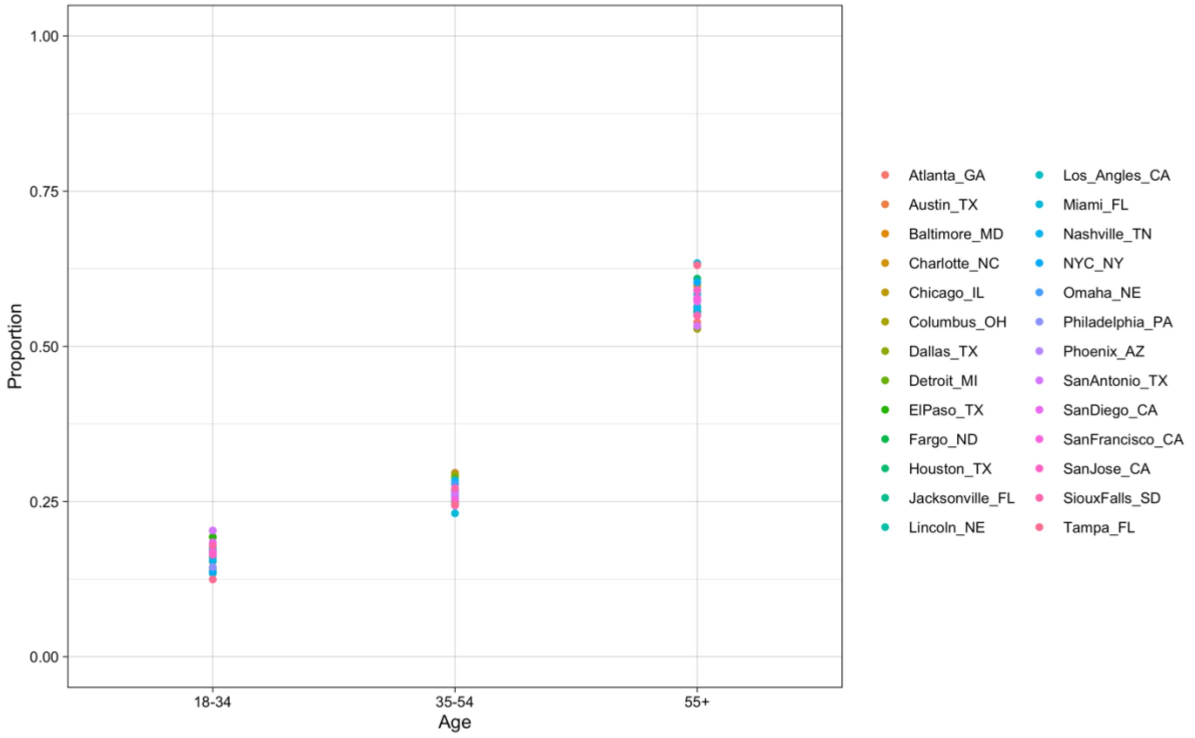


---

Figure S7: Proportion of general US population in each age bracket [61]

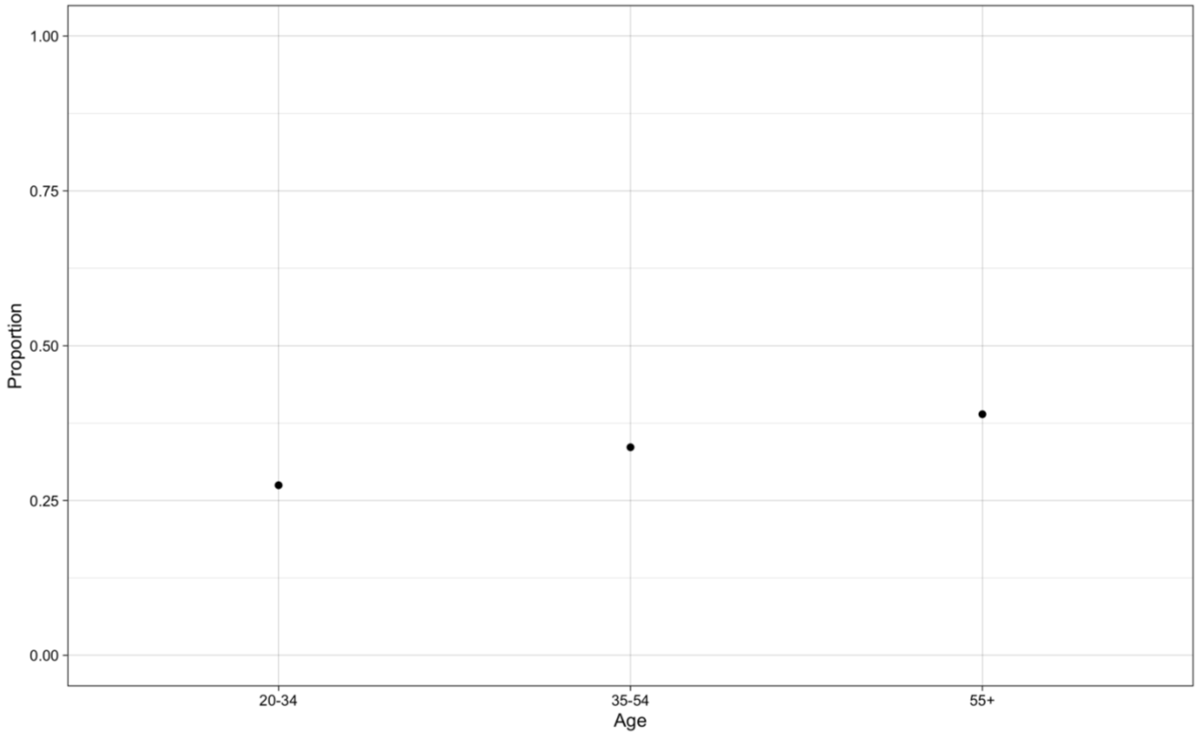

8. Model results with alternative distance quartiles

The results in the main manuscript use distance quartiles based on trips over the whole study period. However, since the frequency of trips of different distances changed different amounts in response to the COVID-19 pandemic, here we present the results of these models when distance quartiles are based on only trips taken in February 2020 (which we took as the baseline period for all analyses). The thresholds for each quartile changed very little and therefore the modelled results were also very similar.

Figure S8: Effects of distance, income, relative case rates, and age on rate of decrease in travel between February 1- April 3 when using distance thresholds defined using trips only during February.

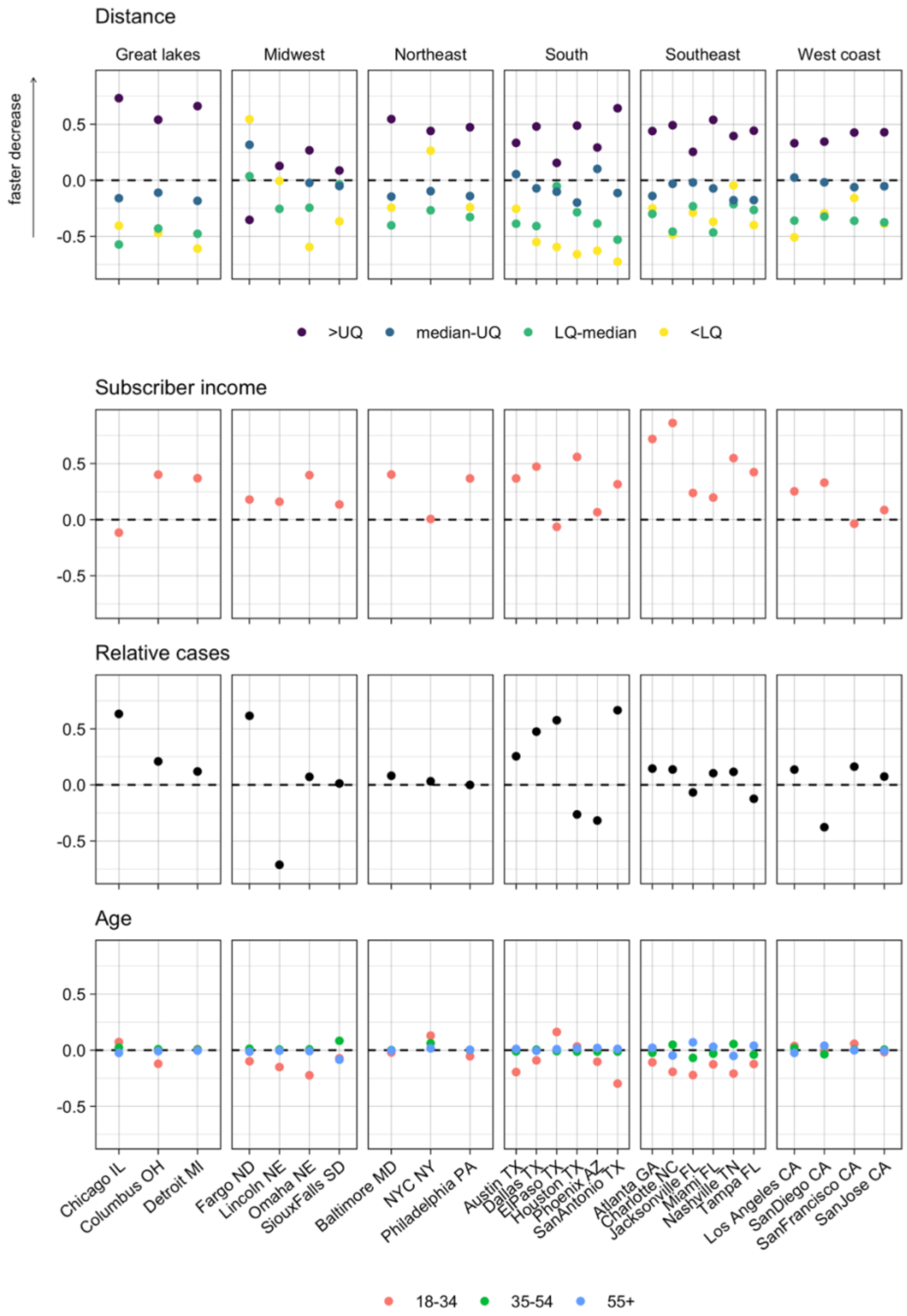

Figure S9: Effects of distance, income and age on trip rates between June 1 – August 31 compared to baseline travel when using distance thresholds defined using trips only during February.

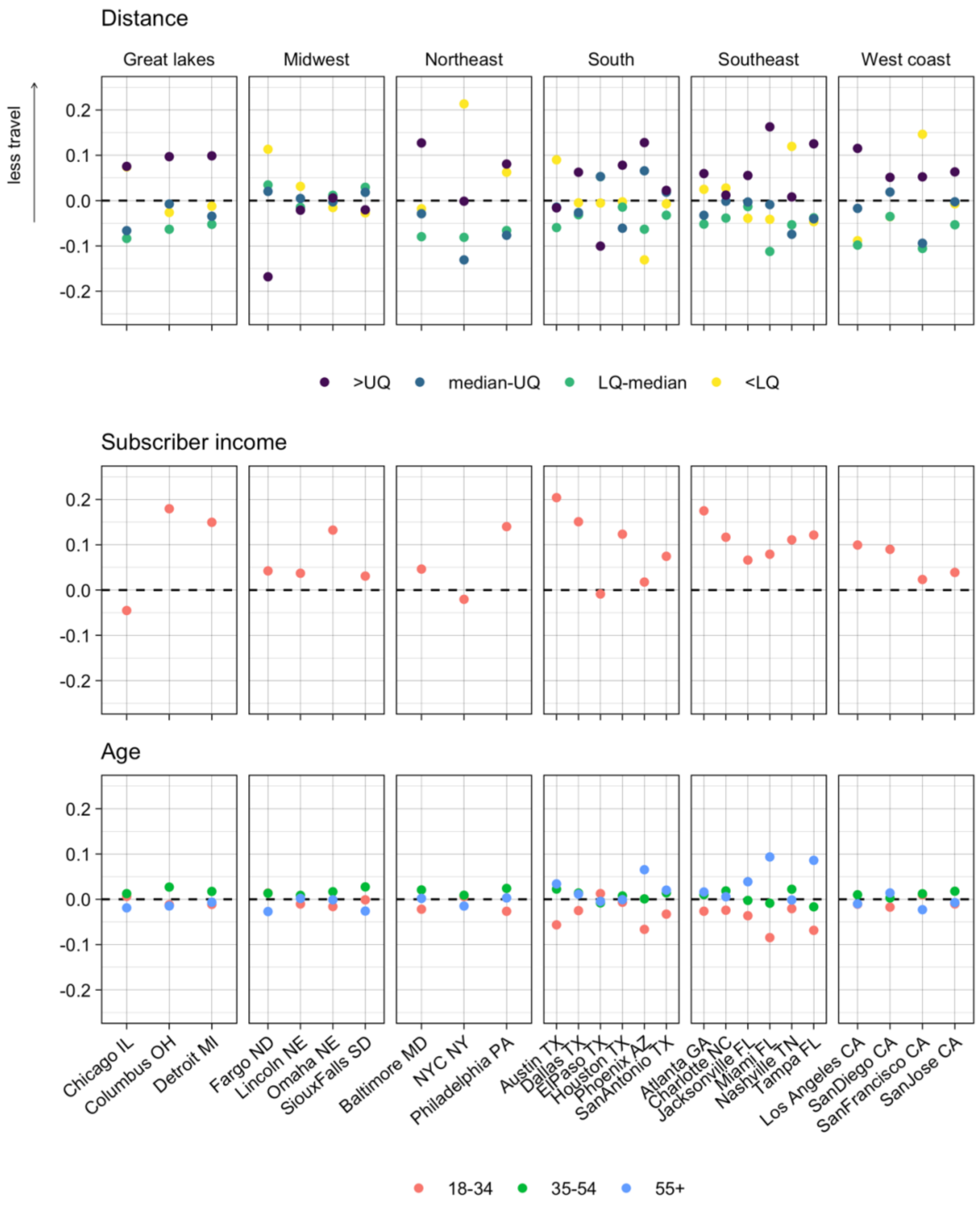

9. Comparison to gravity model

We compare the performance of the original model and where a gravity model was used to model baseline travel. Here we used the Bayesian information criterion (BIC) [46, 47], which is defined as

$$BIC = k\log(n) - 2\log(\hat{L})$$

where k is the number of parameters in the model, n is the number of data points, and L is the maximum likelihood value. Similarly to the Akaike information criterion (AIC), the BIC evaluates model performance based on the likelihood while penalising the number of parameters in the model. This penalisation term is greater in BIC compared to AIC, and therefore we would expect BIC to be more favourable towards the gravity model given the large number of parameters used in the original model. Despite this, BIC values were consistently smaller for the original model compared to the gravity model, suggested the original model performs better.

| City | Original model BIC | Gravity model BIC |
|---|---|---|
| Atlanta | 3840161.2 | 258224101 |
| Austin | 1222027.15 | 64803466.9 |
| Baltimore | 5087018.43 | 261635336 |
| Charlotte | 2067629.99 | 173940690 |
| Chicago | 7706554.13 | 458064822 |
| Columbus | 1938871.43 | 102160615 |
| Dallas | 4518151.76 | 137857475 |
| Detroit | 5297367.31 | 293064915 |
| El Paso | 1195483.35 | 49771459.6 |
| Fargo | 232714.497 | 11924885.5 |
| Houston | 4371970.12 | 245544784 |
| Jacksonville | 719335.198 | 45020737.1 |
| Lincoln | 575325.226 | 74694180.3 |
| Los Angeles | 12295065.1 | 700370925 |
| Miami | 2744751.23 | 159705789 |
| Nashville | 1627853.07 | 591536474 |
| NYC | 43231266.4 | 1235370300 |
| Omaha | 1257801.97 | 333591669 |
| Philadelphia | 6469144.31 | 271283519 |
| Phoenix | 3949295.59 | 1222806194 |
| San Antonio | 1306702.55 | 55910501.3 |
| San Diego | 3323471.85 | 217148993 |
| San Francisco | 5093222.29 | 259613186 |
| San Jose | 3227666.01 | 648414497 |
| Sioux Falls | 257991.936 | 73655260.1 |
| Tampa | 3013279.97 | 175615714 |

### 10. Relationship between travel and NPIs, case rates at the city-level

Figure S10. Relationship between relative travel and NPI stringency (left) and case rates (right) at the city level between (a) March 15 – April 12 2020 and (b) June 1 – August 30 2020. NPI stringency was at the state level. Cities with some significant additional NPIs in place (beyond the state-level policies) between June 1 – August 30 are highlighted in red.
(a) There was little association between travel and either NPI stringency or case rates between March 15 – April 12.
(b) When taking into account cities with additional NPIs, there was some evidence of a weak association between NPI stringency and travel between June 1 – August 30. There was little association between travel and case rates over this period.

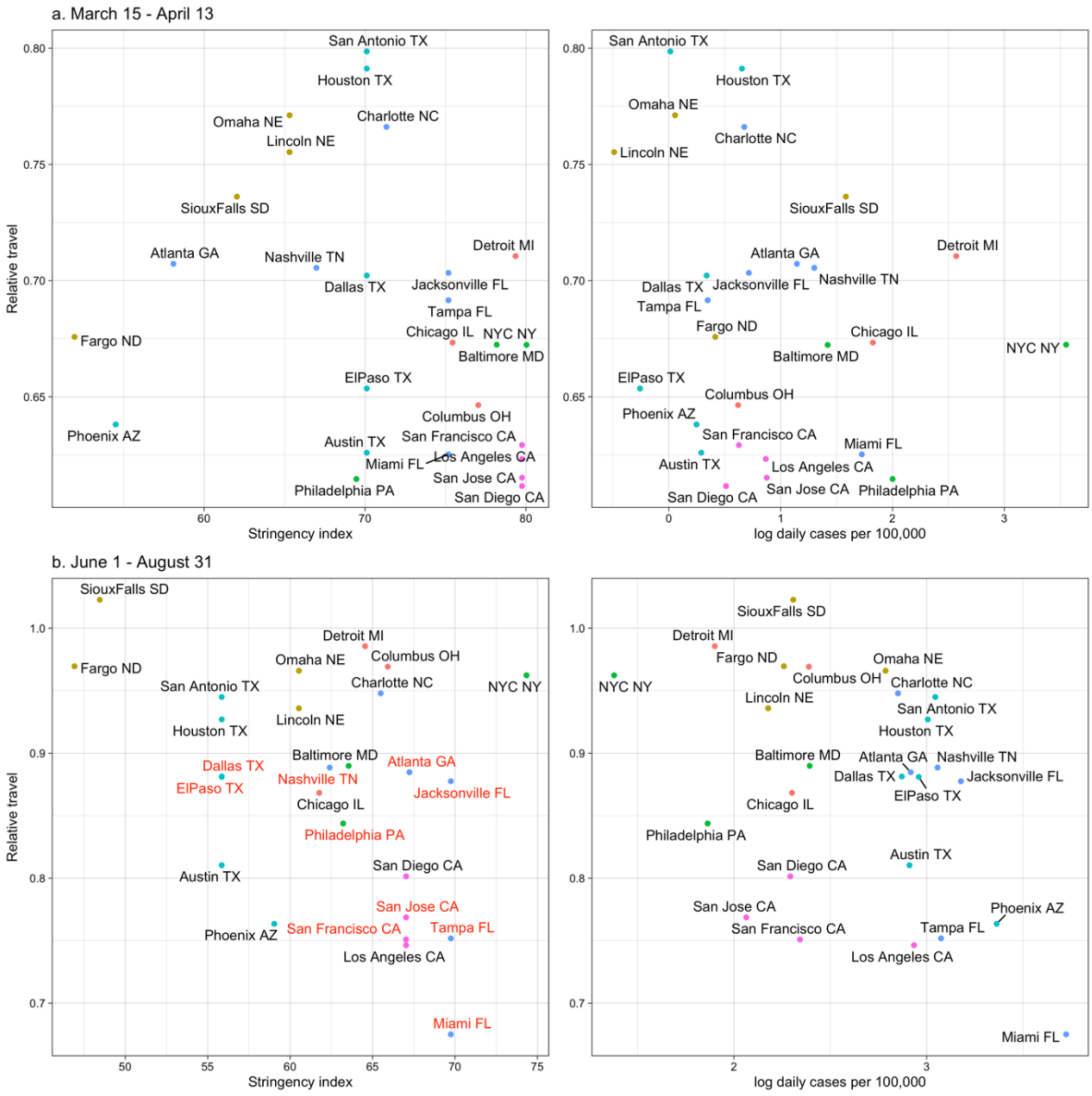
